# Innate immune signaling in the olfactory epithelium reduces odorant receptor levels: modeling transient smell loss in COVID-19 patients

**DOI:** 10.1101/2020.06.14.20131128

**Authors:** Steven Rodriguez, Luxiang Cao, Gregory T. Rickenbacher, Eric G. Benz, Colin Magdamo, Liliana Ramirez Gomez, Eric H. Holbrook, Alefiya D. Albers, Rose Gallagher, M. Brandon Westover, Kyle E. Evans, Daniel J. Tatar, Shibani Mukerji, Ross Zafonte, Edward W Boyer, C. Ron Yu, Mark W. Albers

## Abstract

Post-infectious anosmias typically follow death of olfactory sensory neurons (OSNs) with a months-long recovery phase associated with parosmias. While profound anosmia is the leading symptom associated with COVID-19 infection, many patients regain olfactory function within days to weeks without distortions. Here, we demonstrate that sterile induction of anti-viral type I interferon signaling in the mouse olfactory epithelium is associated with diminished odor discrimination and reduced odor-evoked local field potentials. RNA levels of all class I, class II, and TAAR odorant receptors are markedly reduced in OSNs in a non-cell autonomous manner. We find that people infected with COVID-19 rate odors with lower intensities and have odor discrimination deficits relative to people that tested negative for COVID-19. Taken together, we propose that inflammatory-mediated loss of odorant receptor expression with preserved circuit integrity accounts for the profound anosmia and rapid recovery of olfactory function without parosmias caused by COVID-19.

## Introduction

Subjective reduction of smell (hyposmia) commonly occurs during upper respiratory viral infections (URIs) (*1, 2*), and resolves concomitantly with improvement in rhinorrhea and nasal congestion symptoms. About 5% of patients experience a post-infectious, prolonged olfactory disorder that often recovers over 6 – 12 months with odor training (*3*). By contrast, a much larger proportion of patients infected with the SARS-CoV-2 virus (34 - 65%) self-report anosmia, usually without accompanying rhinorrhea or nasal congestion (*2, 4*). Self-report of smell loss is often unreliable (*5*). Objective smell testing using the 40-item UPSIT smell identification test revealed 98% with olfactory deficits in COVID-19 positive hospitalized inpatients in Iran, in spite of only 34% complaining of loss of smell (*6*). Similarly, 84% of 60 hospitalized inpatients for COVID-19 infection were hyposmic or anosmic using the 12-item Sniffin-Sticks odor identification test whereas only 45% reported subjective loss of smells (*7*). In contrast, in mostly ambulatory patients,the 16-item Sniffin-Sticks odor identification revealed 38% normosmia in COVID positive patients reporting total smell loss (*8*). In many cases, olfactory deficits occur before the onset of other symptoms of a COVID-19 disease or are the only manifestation of the disease (*2, 9*).

Viral induced rhinorrhea, or ‘runny nose’ is one mechanism that may contribute to olfactory dysfunction by preventing odorants from reaching odorant receptors (OR). Infection with SARS-CoV-2, however, is not commonly associated with rhinorrhea or congestion (*2*). Viral killing of olfactory sensory neurons (OSNs), is another mechanism of olfactory dysfunction, and regeneration of the OSNs from stem cells and reintegration of newly differentiated neurons into existing circuits is thought to be responsible for the months-long recovery process. The rate of recovery of olfactory function is another distinguishing feature of COVID-19 associated anosmia relative to other post-infectious olfactory deficits. In a recent longitudinal survey, 80% of patients reported subjective partial or full recovery after 1 week, rather than months as typically described by post-viral smell loss patients (*10*). Together, these distinguishing clinical features (lack of rhinorrhea or congestion, the broad penetrance of hyposmia, and the rapid recovery of olfactory function) suggest that COVID-19 infection induces olfactory loss via a mechanism that is distinct from a neurotoxic effect mediated by other viruses.

Furthermore, the expression of the receptors for SARS-CoV-2, ACE2 and TMPRSS2, on cell-types that are components of the complex cellular composition of the olfactory epithelium, but not directly on OSNs, also suggests a non-cell autonomous model of OSN dysfunction (*11, 12*).

Here, we report that sterile activation of an antiviral signaling cascade in the murine olfactory epithelium interferes with olfactory function by markedly reducing the expression of odorant receptors in OSNs non-cell-autonomously. We previously reported lines of transgenic mice (Nd1 and Nd2) that express genomically encoded cytoplasmic dsRNA in 1% of the cells in the olfactory epithelium. The cdsRNA triggers a sterile type I interferon (IFN-I) innate immune response that spreads to neighboring and connected cells (*13*). The antiviral innate immune response induces hyposmia and a dramatic decrease in odorant receptor RNA levels in both the Nd1 and Nd2 that far exceeds the degree of neuronal loss. Moreover, reversing the IFN-I response by silencing the expression of genomically encoded dsRNA in adult mice affords recovery of OR expression. Our data suggest a model where a robust antiviral innate immune response acts non-cell autonomously to block the expression of functional odorant receptors, which synergizes with OSN cell death to cause olfactory deficits. Based on this model we hypothesize that reduced OR expression would lead to reduced perceived intensity and diminished discriminatory perception in patients infected with SARS-CoV-2. In an initial test of this hypothesis, we characterized olfactory function in non-hospitalized patients that tested positive or negative for the COVID infection by nasopharyngeal RT-PCR for SARS-CoV-2. We find that non-hospitalized patients with COVID-19 infection score intensities of odors significantly lower and discriminate between odors with less acuity relative to patients with negative COVID-19 testing phenotypes that are consistent with a peripheral mechanism of olfactory dysfunction (*14*), and consistent with loss of OR expression. Profound reduction of OR expression in the setting of robust anti-viral innate immune signaling in the olfactory epithelium may resolve the paradox of why SARS-CoV-2 robustly impacts smell function even though OSNs do not express the primary entry receptors for the virus (*11, 12*); this mechanism of OSN dysfunction may also explain why the recovery of profound olfactory function occurs on a short time scale relative to other post-infectious anosmias – days to weeks as opposed to months.

## Results

We generated and characterized the Nd1 and Nd2 mouse models, which express genomically encoded, cytoplasmic dsRNA (cdsRNA). The source of the genomically encoded cdsRNA is their respective transgenes, which integrated into the genome with both intact and inverted copies. The cdsRNA is detected in < 1% of cells in the olfactory epithelium (Fig. 1A). However, this cdsRNA triggers sterile induction of Type I interferon (IFN-I) stimulated genes across the entire olfactory epithelium in a non-cell autonomous fashion. This IFN-I induction mimics a viral infection with an infectivity density of < 1% in the olfactory epithelium (*13*); it also affords the opportunity to investigate the effect of innate immune response separated from any direct toxic effects of the virus. Following purification by FACS sorting to 97% purity (Fig. 1A), mature OSNs profoundly expressed interferon response genes relative to sex-matched littermate controls that did not express the genomically encoded cdsRNA (Fig 1B), (*13*).

**Fig. 1.**
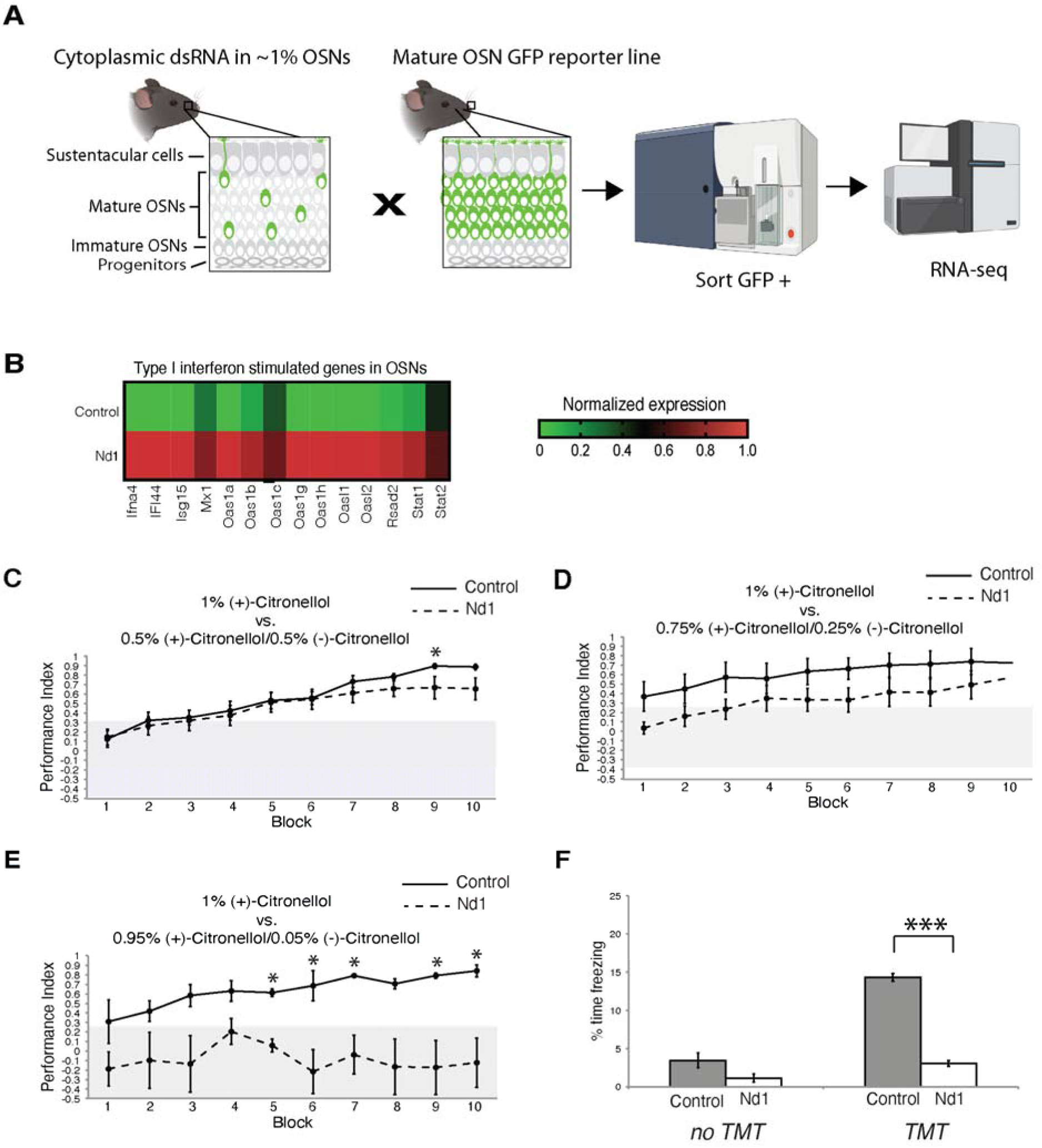
Olfactory deficits in Nd1 mice relative to wild type controls. (A) Overview of experimental design; Cell sorting was used to isolate mature OSNs from progeny of a cross between mice expressing genomically encoded cdsRNA in ∼1% of mature OSNs and mice expressing the green fluorescent protein reporter in all mature OSNs. Messenger RNA was isolated and cDNA was prepared and sequenced from these purified OSNs. **(B)** Heat map of RNA expression of representative IFN-I induced genes expressed in mature OSNs (*5*). **(C)** A cohort of Nd1 (n = 12) mice and littermate controls (n = 13) were trained to discriminate 1% (+)-citronellol from a mixture of (+)- and (–)-citronellol enantiomers at 1:1, **(D)** 3:1 and **(E)** 19:1ratios. The performance index ((correct responses – incorrect responses)/total responses) ±S.E.M. is plotted for each block (20 trials) of a training session (* = p < 0.05). **(F)** Reduced response to odorant with innate behavioral response. Nd1 mice (n = 4) spent less time in a freezing state in the presence of TMT relative to wild-type littermate controls (n = 4) (F_1,6_ = 347, p < 10^−5^ ***). Prior to TMT exposure, the proportion of time in a freezing state was not significantly different (F_1,6_ = 4.22, p < 0.09). Section **(A)** was partially created with BioRender.com.

### Reduced Olfactory Acuity in Nd1 Mice

To determine whether evoking a robust innate immune response in the olfactory epithelium leads to olfactory deficits (*15*), we examined the olfactory acuity of Nd1 mice using two experimental approaches. First, we employed the Slotnick operant conditioning paradigm to assess odorant discrimination (*16*). The task is to distinguish a 1.0% solution of the (+)-citronellol in mineral oil from a racemic mixture of the (+)- and (–)-citronellol enantiomers in mineral oil whose aggregate concentration was 1.0%. Both Nd1 (n = 12) and littermate control (n = 11) mice distinguished a 1:1 ratio of the (+)- and (–)-citronellol enantiomers from 1.0% of the (+)-citronellol enantiomer (Fig. 1C). The performance for Nd1 mice was, however, significantly worse than the control littermates (p < 0.05) when the ratio of the (+): (–)-citronellol enantiomers was increased from 3:1 (Fig. 1D) to 19:1 (Fig. 1E) in the racemic mixture.

In a second experimental approach, the mice were exposed to an odorant isolated from fox feces, 2,5-dihydro-2,4,5-trimethylthiazoline (TMT), a chemical that induces innate aversive responses, including freezing. Mice with a disturbance in the dorsal olfactory bulb do not exhibit a response to TMT (*17*). Nd1 male mice (n = 4; p < 10^−6^) exhibited reduced freezing behavior compared to control male littermates (N=4) when exposed to TMT (Fig. 1F). Together, these data indicate that non-cell autonomous activation of IFN-I response genes in OSNs results in reduced olfactory acuity.

### Reduced Activity of Olfactory Sensory Neurons in Nd1 Mice

Lower odor-evoked activity in the setting of an active innate immune response may account for the reduced olfactory acuity in Nd1 mice (*18*). We assessed of odor-evoked OSN activity by recording local dendritic field potentials generated by odorant stimulation from the surface of the olfactory epithelia of Nd1 and control littermates (*19*). We stimulated three-month-old Nd1 and control littermates with a 1:200 dilution of a saturated vapor of amyl acetate, or a 1:200 dilution of a saturated vapor of pentanal. The local field potentials evoked by either odor were significantly different from the responses to mineral oil control in both Nd1 and control mice (representative traces shown in Fig. 2A and 2B). Similarly, the electroolfactogram responses were markedly different between Nd1 mice (n = 6) and littermate controls (n = 7) for both amyl acetate and pentanal (mean response = 10.43 mv vs. 4.57 mv; p < 10^−100^ and mean response = 11.64 mv vs. 5.22 mv; p < 10^−105^, respectively (Fig. 2C)). Cumulative histogram plots of the distribution of responses for pentanal (Fig. 2D) and amyl acetate (Fig. 2E) revealed a left shift for the Nd1 mice, indicating an effect over the entire population of OSNs. While these results provide evidence of diminished odor-evoked activation in Nd1 mice, the magnitude of the reduced mean responses for both odors (55 and 56%) was greater than the reduction of the steady state levels of OSNs (35%) in Nd1 mice (*13*), suggesting that an additional mechanism beyond OSN death accounts for the reduced mean odor evoked responses.

**Fig. 2.**
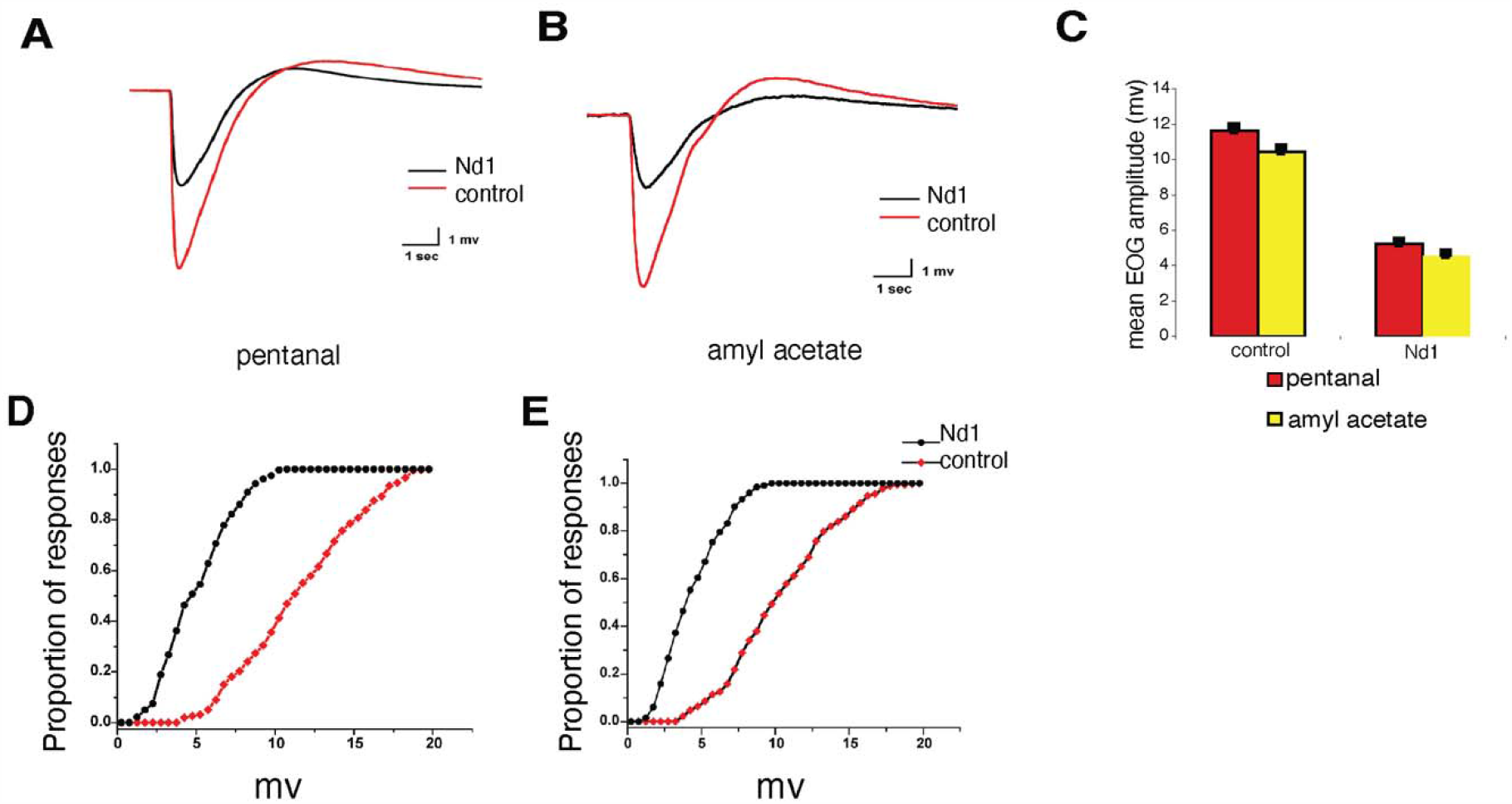
Reduced odor-evoked activity in Nd1 mice. **(A)** Representative local field potential curves from the olfactory sensory epithelia in Nd1 (black) and littermate control (red) in response to the odorant pentanal. **(B)** Representative local field potential curves from the olfactory sensory epithelia in Nd1 (black) and littermate control (red) in response to the odorant amyl acetate. **(C)** Mean EOG amplitudes are plotted for pentanal and amyl acetate for Nd1 and littermate control mice (p < 10^−100^). **(D)** Cumulative histogram of electroolfactogram responses of Nd1 and littermate control mice to 1:200 dilution of pentanal. **(E)** Cumulative histogram of electroolfactogram responses of Nd1 and littermate control mice to 1:200 dilution of amyl acetate.

### Marked Reduction of Expression of Multiple Classes of Odorant Receptors in Response to Anti-Viral Signaling

We tested the hypothesis that reduced odorant receptor expression, coincident with the partial loss of OSNs, could account for the loss of olfactory acuity in Nd1 mice. Although murine OSNs carry greater than 1000 genes for distinct odorant receptors (OR), each OSN expresses only a single odorant receptor. We examined members of three different classes of ORs; class I, class II, and TAAR receptor families (*20, 21*). We used RNA *in situ* hybridization on coronal sections of olfactory epithelia to detect representative OR members from each class. We found a significant reduction (70-92%, p >0.0001 for all ORs examined) in the number of OSNs expressing representatives from all three classes of ORs (Fig. 4A-4D).

We next examined the same three class representative ORs in Nd2, a second transgenic mouse line with a similarly genomically encoded trigger of robust innate immune activation (*13*). We also found a robust reduction in odorant receptors in Nd2 (Fig. S1; 75-98%, p >0.0001 for all ORs examined). Using qPCR we independently confirm the reduced expression of 32 different ORs from all three classes (Fig. 3E). Next we performed a metanalysis of RNA-seq data from purified mature OSNs to determine if OR loss is widespread across OSNs we analyzed the expression of odorant receptors (*13*). We detected a significant reduction in the expression of 933 odorant receptors, TAAR receptors, and OR pseudogenes (Fig. 4A). Together these data show that odorant receptor expression is reduced across most OSNs throughout the olfactory epithelium in response to sterile innate immune activation.

**Fig. 3.**
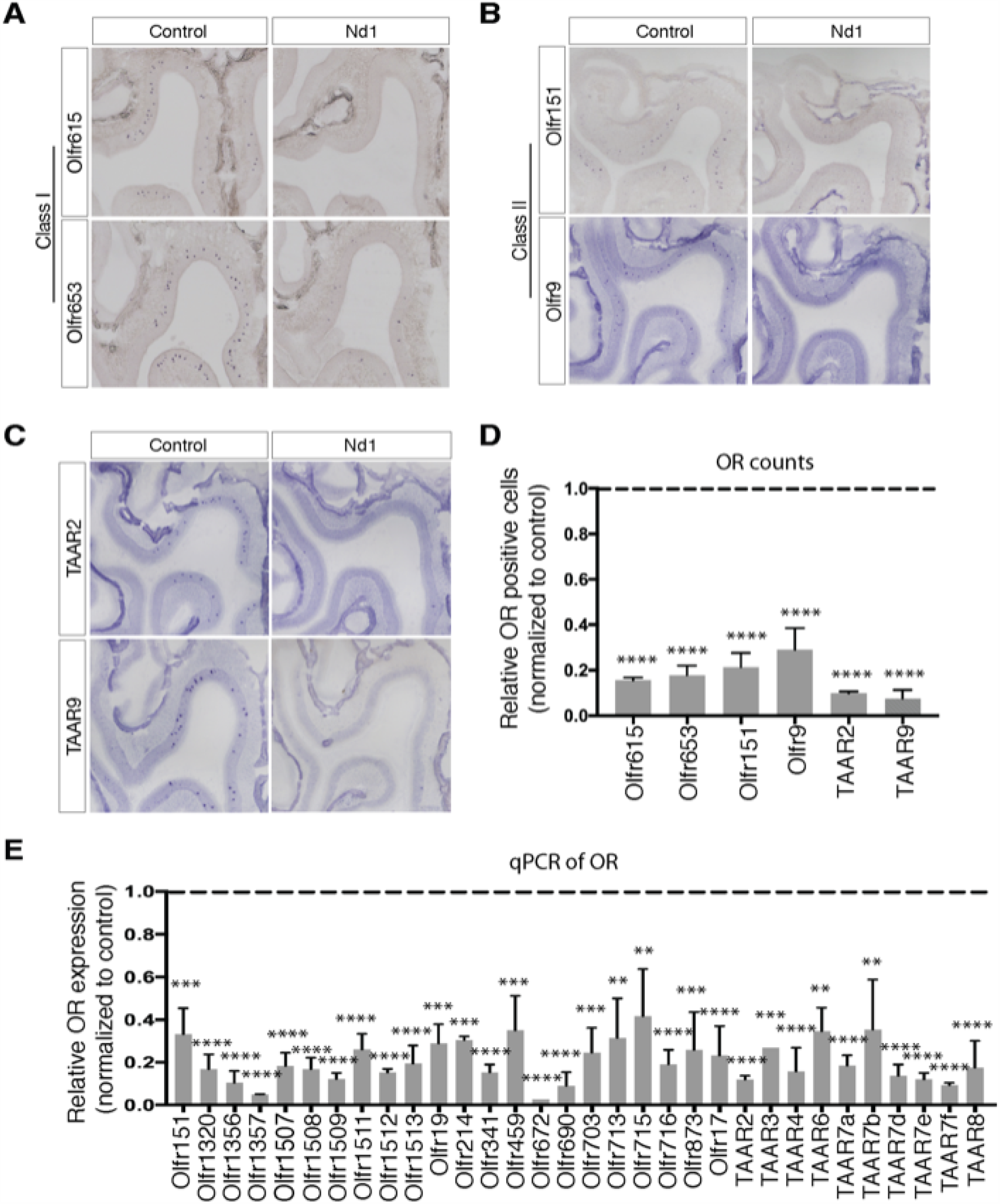
Markedly less OR expression in OSNs in Nd1 mice. **(A)** Representative regions of olfactory epithelium detected by RNA *in situ* hybridization with probes to Class I OR representatives Olfr615 and Olfr653 in Nd1 and a control littermate. **(B)** Representative regions of olfactory epithelium detected by RNA *in situ* hybridization with probes to Class II OR representatives Olfr151 and Olfr9 in Nd1 and a control littermate. **(C)** Representative regions of olfactory epithelium detected by RNA *in situ* hybridization with probes to TAAR receptor representatives TAAR2 and TAAR9 in Nd1 and a control littermate. **(D)** Summary of total counts of littermate pairs of control and Nd1 mice, revealing a marked reduction of all representative ORs, which is statistically significant (****p ≤ 0.0001; n=3 mice). **(E)** Summary of RNA expression for 39 representative ORs by qPCR (**p ≤ 0.01, ***p ≤ 0.001, ****p ≤ 0.0001).

**Fig. 4.**
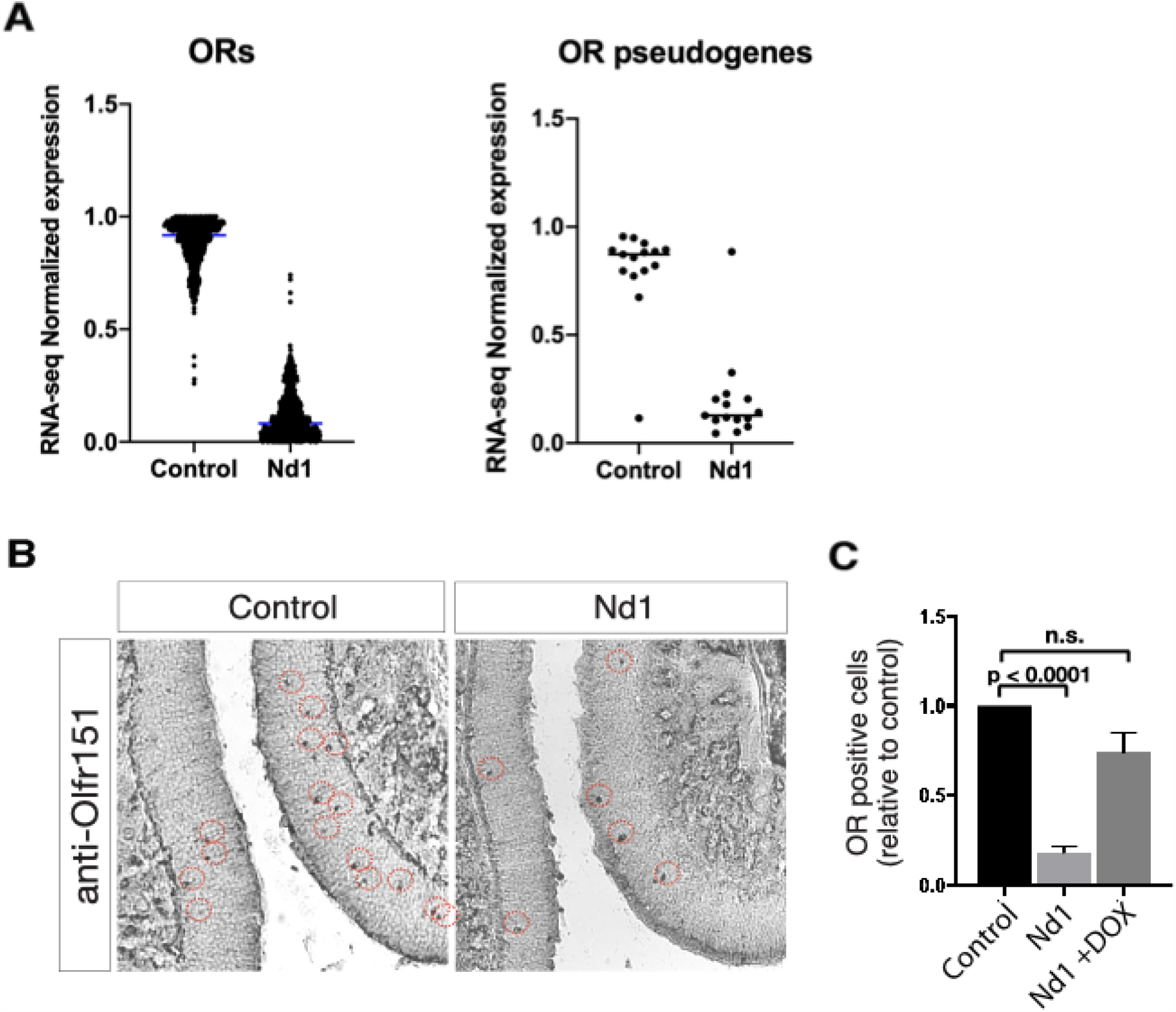
Global repression of OR transcription and reversal of OR repression by suppression of transgene expression. **(A)** Relative RNA levels of OR genes (n=933), and OR pseudogenes (n=15), expressed in mature OSNs by RNA-seq of purified OSNs by FACS (*13*). **(B)** Representative regions of olfactory epithelium immunostained with an anti-Olfr151 antibody in a control and Nd1 littermates. **(C)** Quantification of Olfr151 receptors in Nd1 and in Nd1 after suppression of transgene expression for 2 months relative to littermate control mice on doxycycline.

We explored the consequences of repression of IFN-I signaling by silencing transgene expression by feeding Nd1 mice doxycline to induce the tetracycline repressor engineered into this mouse line. Extinguishing the viral-mimetic expression in OSNs for 2 months in young adult mice reduced the number of cleaved caspase 3 positive cells in the olfactory epithelium to levels similar to those observed in control animals (*13*). Using immunohistochemistry, we found that OSNs expressing the Olfr151 recovered to levels that were not statistically different than control littermates in Nd1 (Fig. 4C) but were significantly different to Nd1 mice that were not fed doxycycline (Fig. 4B and 4C; p < 0.0001). These observations indicate that resolution of anti-viral inflammatory response in the olfactory epithelium results in resumption of OR expression, and that the lower gene expression levels of ORs was not due to aberrant development.

### Reduced subjective odor intensity and odor discrimination in COVID-19 infected patients relative to individuals testing negative for COVID-19

Based on our mouse studies, we hypothesized that COVID-19 infection of respiratory epithelial and sustentacular cells leads to local innate immune activation (*11, 12*), even in the absence of constitutional symptoms to reduce OR expression in OSNs. To test this hypothesis in humans, we repurposed our olfactory battery developed for early diagnosis of cognitive impairment (*22*) into a smell test for probing olfactory function in people with a possible or definite exposure to SARS-CoV-2 virus. Reviewing our data collected on control participants from ages 20 - 89, we selected the three complex odor mixtures with the highest percentage of accurate responses on the odor percept identification test (80%, 85%, and 90%, respectively). The COVID smell test battery consists of three sequential tests: 1) rate the intensity of each odor, where participants describe the potency of a particular smell; 2) odor percept identification, where participants select from four choices the name that best describes the odor they remember smelling; and 3) discrimination, where the participant smells three odors consecutively and then selects which one of the three odors is different. We presented odors as peel and sniff labels arrayed on a card and recorded responses on web-based app (Fig. 5A).

**Fig. 5.**
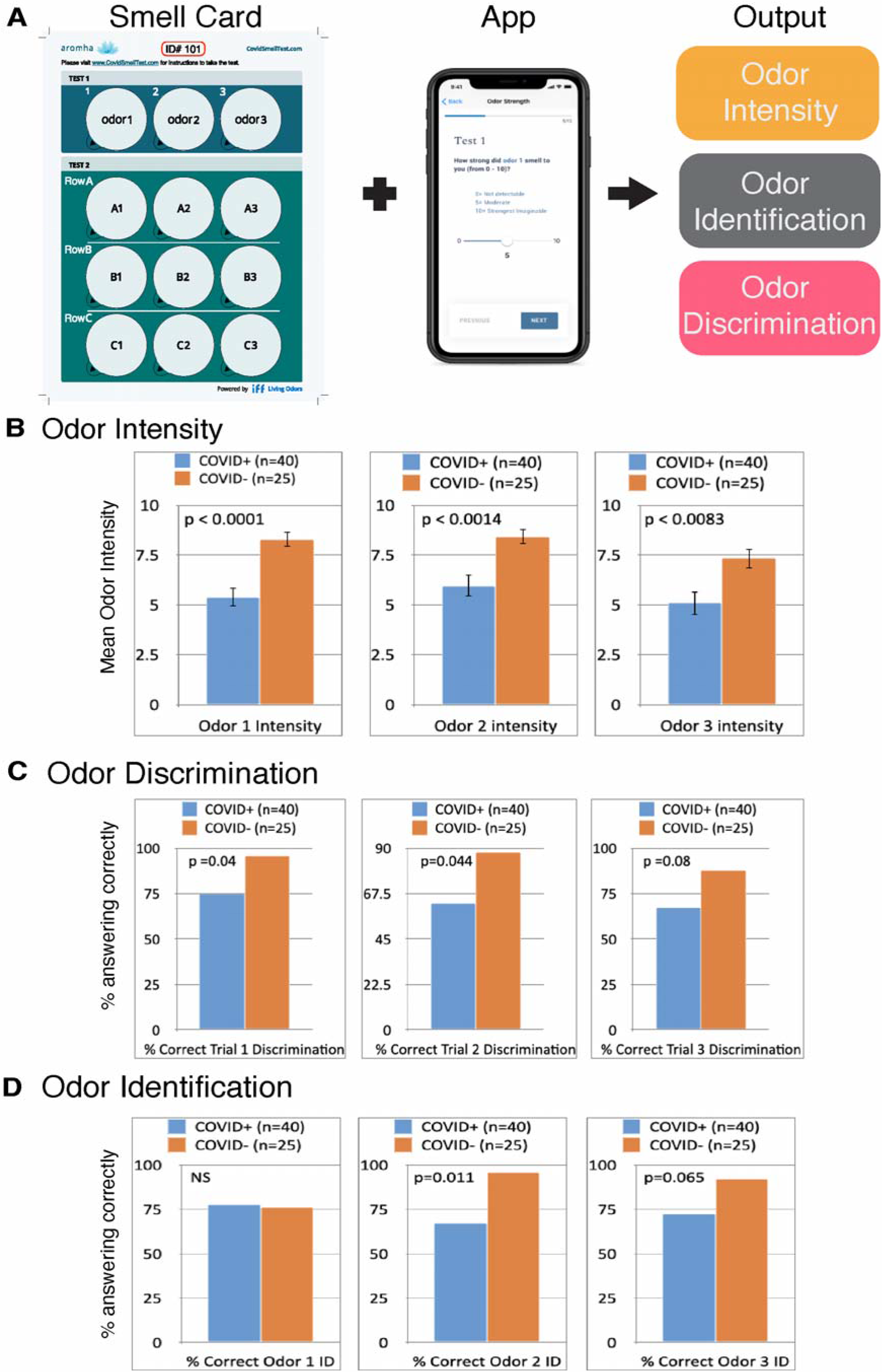
Reduced odor intensity ratings and reduced odor discrimination in patients and healthcare workers infected with COVID-19 virus relative to patients and healthcare workers who tested negative for COVID-19 by the by the nasopharyngeal RT-PCR assay. **(A)** Overview of the COVID Smell Test. **(B)** Odor intensity ratings were significantly reduced in COVID-19 patients relative to participants who tested negative for COVID-19. **(C)** Odor discrimination scores were significantly reduced in COVID-19 patients relative to participants who tested negative for COVID-19. **(D)** Odor percept identification scores were not significantly reduced in COVID-19 patients relative to participants who tested negative for COVID-19. ***p ≤ 0.001; ** p < 0.01; * p < 0.05

Patients with documented SARS-CoV-2 by nasopharyngeal swab RT-PCR tests (n = 40) and patients with one or more negative SARS-CoV-2 RT PCR test results (n =25) were recruited to take the COVID Smell Test. Consistent with previous reports of symptoms, subjective complaints of smell loss were greater in the COVID-19 patients, but the distribution of age, sex, time between the RT-PCR assay and smell test were not statistically different between the two cohorts (Table S1). COVID-19 patients demonstrated reduced subjective intensity ratings for all three odors (Fig. 5B), discriminated two of three odor combinations significantly worse – with a trend in the third combination - compared to patients with negative SARS-CoV-2 test results (Fig. 5C). By contrast, only one of the three items of the odor percept identification test was significantly different between these two groups with a trend in another odor (Fig. 5D). Together, the reduced intensities and difficulty with discriminating 3 different presented odors is consistent with a peripheral mechanism of olfactory dysfunction (*23*).

Another distinguishing feature of anosmia in the setting of COVID-19 is the recovery of smell function within days to weeks (*10*) – as opposed to months (*3*)- in many patients without distortions (parosmias). These differences are illustrated in a case of a 60 year old female with no significant previous health history who developed an upper respiratory illness with nasal congestion for 2 weeks and was left with a persistent anosmia for 8 months (from 4/2019 until late 12/2019). Her initial recovery of smell function was notable for identical odor-evoked percepts - independent of the nature of the odor - that was distorted and unpleasant. Over the next 3 months, her smell function improved about 75% of her subjective baseline – distinguishing between different red wines – when she suddenly lost her sense of smell in mid-March 2020 without nasal congestion. Her other two family members became ill with an upper respiratory illness, and her daughter also lost her sense of smell transiently. This second bout of anosmia was preceded by hosting a visitor from Spain. However, her sense of smell began to recover in 3 days and was not associated with distortions – described as faint, accurate perceptions and within 4 weeks she had regained more subjective smell function than she had experienced before the second bout of anosmia. This case vividly illustrates differences between post-infectious anosmia that is thought to be caused by a neurotoxicity of OSNs – her 2019 bout of anosmia – where recovery of smell function requires the regeneration of OSNs from stem cells, and reintegration of nascent OSNs into existing neural circuits. Transient parosmias may represent the transition from the initial synaptic connections to refined integration mediated by synaptic plasticity. In her 2020 bout of anosmia, which was equally profound, the olfactory neural circuit appears to be rendered transiently dormant.

## Discussion

In this study we show that non-cell autonomous induction of a sterile innate immune response across the murine olfactory epithelium reduced olfactory acuity, reduced odor-evoked potentials, and dramatically reduced levels of odorant receptor RNA and protein in olfactory sensory neurons (Fig. 6). We previously demonstrated a 35% loss of OSNs in these mouse lines (*13*). Both surgical and genetic studies demonstrated that 5% of the baseline OSN population is sufficient for nearly normal smell function (*24, 25*). Therefore, we posit that a combination of death of a subset of the OSN population and the repression of odorant receptor transcription and protein expression cause the olfactory deficits in the Nd1 mouse line.

**Fig. 6.**
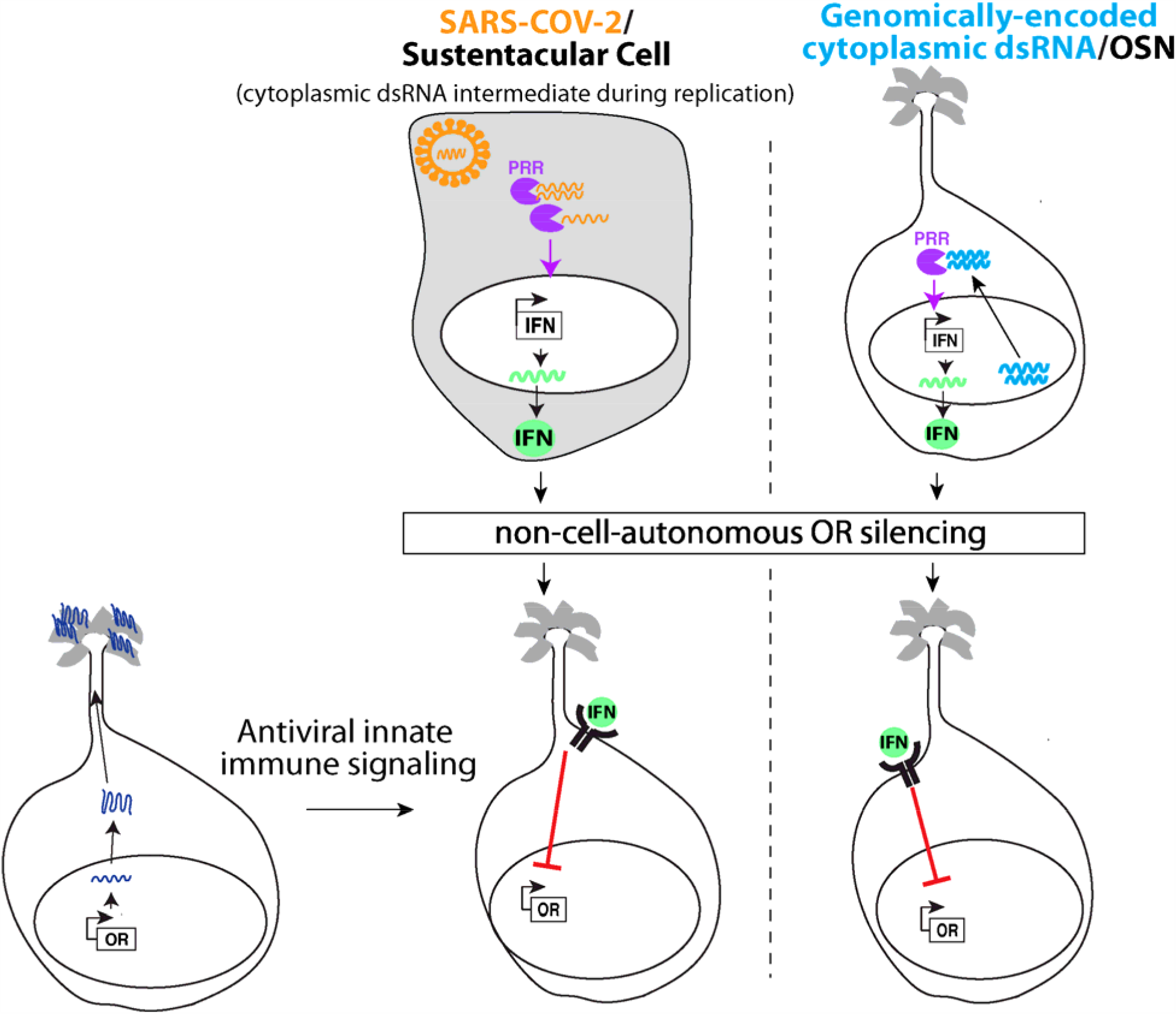
Model for non-cell-autonomous innate immune inhibition of odorant receptor expression. All cells in the olfactory epithelium, including mature OSNs express pattern-recognition receptors (PRR) that can detect cytoplasmic or endosomal nucleic acids. For example, the positive RNA single stranded genome of Coronaviruses, or its double-stranded RNA intermediates, can be detected by PRRs. PRRs then activate the transcription of interferon (IFNs) cytokines, and other cytokines and chemokines, which are secreted from infected cells. IFNs then trigger the activation of a host of genes that define the type I interferon antiviral innate immune response. In Nd1 and Nd2 transgenic mouse lines, ∼1% of OSNs activate PRRs, and the other 99%, including non-neuronal cells respond to secreted IFN. IFN-I then leads to repression of OR expression. During COVID-19, even though OSNs do not express the main entry receptors for SARS-CoV-2, they can still detect IFNs, other cytokines and chemokines, from sustentacular or other neighboring cells, which activate a robust IFN-I response in OSNs, leading to repression of OR transcription.

We postulate that the profound and common olfactory deficits in the setting of COVID-19 infection may operate via a similar non-cell autonomous innate immune mediated mechanism of OSN dysfunction. As a positive strand RNA virus, the replication cycle of SARS-CoV-2 will include cytoplasmic dsRNA intermediates (*26*), which are pathogen associated molecular patterns that can elicit a robust innate immune response. Two percent or more sustentacular cells in the olfactory epithelium, but not OSNs, express ACE2 and TMPRSS2 entry receptors necessary for robust infection by SARS-CoV-2 (*11, 12*) (Fig. 6). Based on observations in our Nd1 and Nd2 mouse models, viral infection of less than 1% of cells in the epithelium is sufficient to activate innate immune signaling in nearly all OSNs across the olfactory epithelium. We hypothesize that IFNs, or other cytokines that are secreted by infected sustentacular cells can activate an antiviral response within OSNs and suppress OR expression. This model is consistent with previous work that demonstrated selective expression of interferon-gamma exclusively in sustentacular cells can induce anosmia without damage to the neuroepithelia (*27*). In addition, cultured OSNs from mice intranasally inoculated with Sendai virus, a respiratory virus, which triggers a robust type I interferon response (*28*), are less responsive to odorant mixtures (*18*), but the expression patterns of ORs in OSNs were not formally evaluated.

While our data and the data from these previous studies demonstrate that interferon signaling correlates with OSN dysfunction, the precise nature of the innate immune response elicited by COVID-19 in the olfactory epithelium remains to be determined. A hyperactive innate immune response triggered by SARS-CoV-2 is hypothesized to mediate the respiratory failure in patients with COVID-19 (*29, 30*). However, recent studies indicated that respiratory cells infected with SARS-CoV-2 might elicit a weaker Type I interferon response relative to the chemokine response (*31, 32*). Two recent studies indicate that the presence of smell loss is associated with reduced risk of developing moderate to severe clinical stages of COVID-19 infection (*33*). The magnitude of the interferon response in each individual might account for inverse correlation between the smell loss and severe respiratory illness. Additionally, we also cannot rule out release of dsRNA into the extracellular milieu, which could trigger TLR3 signaling leading to olfactory dysfunction (*34*).

The mechanism by which increased interferon signaling disrupts OR expression remains to be fully defined. The choice OR allelic selection begins in immature OSNs (which do not express the cdsRNA in Nd1 and Nd2 mice), and immature neurons robustly express interferon response genes (*13*). The finding that OSNs in Nd1 and Nd2 express cdsRNA in mature OSNs does not preclude IFN-I signaling originating in other cells types from silencing OR expression in a non-cell-autonomous way (Fig. 6). The translation of a functional OR triggers the phosphorylation of EIF2A (Eukaryotic translation initiation factor 2A) by PERK (protein kinase R (PKR)-like endoplasmic reticulum kinase) (*35*). Activation of the dsRNA sensor PKR (protein kinase RNA activated) phosphorylates the same serine 51 residue on EIF2A as PERK (*36*). Activation of PKR requires dimerization, which occurs by increasing its local concentration of PKR on dsRNA templates or by increased expression via IFN-I signaling (*37*), and we have previously observed increased PKR levels in OSNs in Nd1 mice (*13*). We speculate that increased IFN-I activity could trigger the premature phosphorylation of EIF2A leading to silencing of all OR transcription prior to the production of a functional OR. Second, lysine methyltransferases are interferon response genes, which place three methyl groups on Lysine-9 of Histone 3 to produce H3K9me3, a marker for transcriptionally silent olfactory receptors (*38, 39*) (*40*). Therefore, an alternate, but not mutually exclusive hypothesis, is that IFN-I induction might lead to premature formation of H3K9me3 at OR loci prior to the selection of a functional OR.

Lastly, the nuclear organization of OR loci into specific domains within OSNs is an important feature for the selection of a single OR gene, while silencing all other ORs (*41*). The IFN-I antiviral response requires a dramatic change in the expression of genes across the entire genome (cite PMID 18614013; PDF 1198). For example, 10% (>1000 transcripts) of detectable transcripts are differentially expressed in Nd1 relative to controls (*3*). Therefore, the nuclear architecture required for the induction of IFN-I induced genes (*42*) may not be compatible with OR expression. Future studies will investigate these mechanisms in our mouse model and characterize the expression of human OR genes in human olfactory epithelium from patients with documented COVID-19 infection and objective smell dysfunction. If this model is confirmed in human olfactory epithelia of patients with COVID-19 infection, then Nd1 and Nd2 transgenic mice provide a model to characterize the efficacy of therapies to reduce innate immune-induced toxicity in the olfactory epithelium that is separate from direct toxicity mediated by pathogenic viruses.

In COVID-19 infected patients, the olfactory phenotypes of diminished perceived intensity and diminished discrimination in the participants with documented COVID-19 infection are consistent with peripheral olfactory deficit (*23*), such as downregulation of ORs, as seen in the Nd1 mouse, or disturbed extracellular milieu due to sustentacular cell dysfunction (*11*). The weaker association of odor percept identification responses to COVID-19 status relative to the discrimination deficit is distinct from patients with Alzheimer’s disease (*22, 43*) and or Parkinson’s disease (*44*), where olfactory dysfunction is likely a central (brain) disorder. Recent reports using highly validated smell identification batteries (*6, 7*) demonstrated deficits in hospitalized COVID+ patients, but they did not access subjective intensity or discrimination. Some differences in our study that may account for the weaker association between smell identification and COVID status include a smaller sample size (*6, 7*) and 3 items in our battery relative to 16 and 40 items in the Sniffin’ Sticks and UPSIT batteries, respectively, different intensities of the odors, the composition of the odors presented, non-hospitalized patients in our study. Another key differences between our identification battery and the other batteries include that participants see the choices of name before smelling the odor vs. after smelling the odor in our odor percept identification test..

Anosmia in COVID-19 has been reported to recover within two weeks after other symptoms dissipate, which is too rapid for regeneration and differentiation of OSNs followed by axon extension and reintegration into the olfactory bulb circuits in humans (*45, 46*). In addition, the absence of parosmias in the course of recovery suggests that peripheral olfactory neural circuit remains intact and is rendered dormant in many cases of COVID-19 infection. We postulate that resolution of the innate immune signaling evoked by the SARS-CoV-2 could restore expression of olfactory receptors in dormant OSNs and restore the physiologic milieu around OSNs to a physiologic state to afford recovery of smell function that is weaker but not distorted. If the olfactory neural circuit remains intact, then the input into the olfactory bulb can be processed equivalently by central neural circuits in the pre-infectious state. Future work will characterize the utility of objective smell testing combined with symptom tracking and contact tracing to detect COVID-19 infection in asymptomatic individuals and to characterize the temporal correlation of return of olfactory function documented by longitudinal testing with the emergence of markers of immunity to COVID-19.

In summary, we propose a model where inflammatory-mediated loss of odorant receptor expression with preserved circuit integrity contributes to the profound anosmia and rapid recovery of olfactory function without parosmias caused by COVID-19. If this model withstands further investigation, self-administered, objective smell testing may serve as a biomarker for the inflammatory status in the olfactory epithelium evoked by viral infections, including COVID-19 and other coronaviridae.

## Data Availability

All data for this study is available with this submission, except raw clinical data with personal health information, or at Synapse (mouse RNA-seq data-syn22161010).

https://www.synapse.org/#!Synapse:syn22161010

## Acknowledgements

The authors thank Richard Axel for critically reading the manuscript for support as some of the experiments were conducted in his lab, the programming team at the ADK group for development of the COVID Smell Test app, International Flavors and Fragrances for donating odors and insights, MFR Samplings for donating odor labels and printing smell cards, the research staff supporting COVID research at Mass General Brigham, Michael Leone, Albert Hung, Marcelo Matiello, Bradley T. Hyman, Lee Schwamm, and Merit Cudkowicz at MGH for advice, and all the patients and their families for participating in this research.

## Author Contributions

Conceptualization, S.R and M.W.A; Methodology, S.R., C. M., D. J. T., and M.W.A.; Data Generation: S.R., L.C, G.T.R, E.B, K.E., C.R.Y., M.W.A.; Clinical Data Collection: C.M., L.R.G., E.H.H., A.D.A., R.G., M.B.W., S. M., R. Z., E. W. B., M.W.A. Data Analysis, S.R., L.C., E.B., C.M., K.E., C.R.Y., A.D.A., M.W.A.; Resources, S.R., M.W.A; Data Curation, S.R., M.W.A; Writing - Original Draft, S.R and M.W.A. All authors revised the paper.

## Materials and Methods

### Animals

All mouse work was in accordance with protocols approved by the Institutional Animal Care and Use Committee (IACUC) of Massachusetts General Hospital. All transgenic mouse lines have been previously described (*5, 24*). Doxycycline was administered at 40 mg/g in food (Bioserve).

### Humans

All human research was in accordance with the Institution Review Board (IRB) at Mass General Brigham. Patients presenting to the Respiratory Illness Clinics for evaluation at MGH and BWH, patients in an isolation hotel in Revere, MA who were recently hospitalized for COVID-19 but could not return home, and healthcare workers at MGH and BWH were recruited with flyers in English and Spanish.

### Mouse Behavior

The enantiomeric discrimination assays were based on the Slotnick paradigm (*16*). All odorant enantiomers were the highest grade available (Sigma-Aldrich) and diluted in light mineral oil. Nd1 (3 – 9 month old (n = 12) and littermate controls (n = 13) were water restricted (approximately 1 – 1.5 mL per day) for 7 days prior to training as well as during the training and testing. Body weights were followed to ensure that they did not fall below 85% of their baseline weight. Training and testing were performed on an 8-channel liquid dilution olfactometer (Knosys, Lutz, FL). Mice were first trained to lick from the sampling tube, with increasing durations between odor presentation and water reward. The program was repeated until each mouse could complete 200 trials; learning was assessed by a decrease in number of “incorrect” licks and a constant number of “correct” licks. A successful training or testing session consisted of a performance index (the ratio of “correct” licks minus “incorrect” licks over the total licks) to be greater than 0.25. After successful completion of the initial training program, mice were trained to discriminate 2% ethyl acetate in light mineral oil and air. Then the last training session required mice to discriminate 2% ethyl acetate in light mineral oil and 0.1% ß-citronellol in light mineral oil. For testing, the following odor pairs were presented sequentially: 1:1 ratio (0.1% (+)-ß-citronellol vs. a mixture of enantiomers (0.05%(–)- / 0.05%(+)- ß-citronellol in light mineral oil for ORCA and (1.0 % (+)-ß-citronellol vs. a mixture of enantiomers (0.5%(–)- / 0.5%(+)- ß-citronellol in light mineral oil for ND1); 1:3 ratio (0.1%(+) ß-citronellol vs. 0.075%(+)/0.025%(–) ß- citronellol for ORCA and 1%(+) ß-citronellol vs. 0.75%(+)/0.25%(–) ß-citronellol for ND1; and 1:19 ratio (0.1%(+) ß-citronellol vs. 0.095%(+)/0.005%(–) ß-citronellol for ORCA and 1%(+) ß-citronellol vs. 0.95%(+)/0.05%(–) ß-citronellol for ND1. The performance index was calculated from the raw data, and each block was analyzed using an unpaired t-test.

The TMT assay was derived from the curtain assay (*2*). Briefly, approximately 2 month old male ND1 (n = 4) and littermate control (n= 4) mice were habituated individually to a clean mouse cage separated by a parafilm curtain partition that divides the arena into two compartments – one twice as large as the other – for 10 min. This habituation was repeated immediately in a distinct clean cage with the same partition. Then the mice were placed into a new clean cage with the same partition in a chemical hood, and the subsequent 10-minute interval was recorded. After 7 minutes, two pieces of filter paper were taped to opposing walls of the cage. The paper in small compartment had absorbed 20 µL TMT (Contech), and the filter paper in the large compartment had adsorbed 20 µL water. The video was analyzed for freezing, and the observer was blinded to the genotype. These data are analyzed using a single factor ANOVA (% of time freezing as a function of genotype).

### Electroolfactogram

Local field potentials (EOGs) were recorded from the surface of intact olfactory sensory epithelia as described previously (*19*). Amyl acetate and pentanal were applied at 1:200 of saturated vapor (1:100 dilution in mineral oil and 1:2 dilution in the air stream).

### Immunohistochemistry and in situ Hybridization

Immunohistochemistry was performed as previously described (*5*). guinea pig anti-Olfr151 (1:1000). RNA *in situ* hybridization was performed on 20µm thin sections of frozen olfactory epithelia as previously described (*<u>1</u>*). PCR was used to amplify OR sequences from oligo-dT primed cDNA libraries made from whole olfactory epithelia, cloned into vectors, and transcribed into Digoxigenin labeled probes as previously described (*47*).

### qPCR

We used the iQ Sybr Green Supermix (catalog # 170-8880, Bio-Rad, Hercules, CA) to perform qPCR in technical triplicates on an iCycler (Bio-Rad, Hercules, CA) with a tm =60 degrees. At least 2-4 biological replicates were performed. Q-PCR primers were designed using PrimerBank (*8*), TAARS as in used in (*48*), and were only used for analysis if they had an efficiency of at least 90%.

### Human olfactory testing

Participants were given a uniquely numbered smell card (Fig. 5A) and instructed to visit the website (www.covidsmelltest.com), which described the study in English or Spanish with the option of watching an introductory video in English and in Spanish. It then linked participants to the e-consent framework on REDcap, available in both English or Spanish, where they entered their name, their birthdate and their unique smell card number into REDcap. Participants self-administered the COVID Smell Test using their own phone, tablet, or computer by following instructions to peel labels on the smell card and sample the odors (Fig. 5). Participants were asked to rate the intensity of each odor and to undergo tests for odor identification (Test 1) from four choices and odor discrimination (Test 2) where participants selected the present odor that differed from the other two odors.

Identifiable information was only accessed by research staff through the REDCap project and was utilized to access participant medical record number and their electronic health record to collect relevant medical variables and the timing and results from the COVID testing and the type of test used. These results were combined with the ID# on the smell card for analysis by the statistician as deidentified data.

### Statistical Analysis

One-way analysis of variance and unpaired t-tests were used, as appropriate, for the mouse studies. Kruskal-Wallis test, Fisher’s exact T-test, paired and unpaired t-tests were used to analyze the human olfactory responses. Each statistical test was performed as described in the methods and figures legends using SAS, Microsoft Excel, or Apple Numbers.

---

**Fig. S1.**
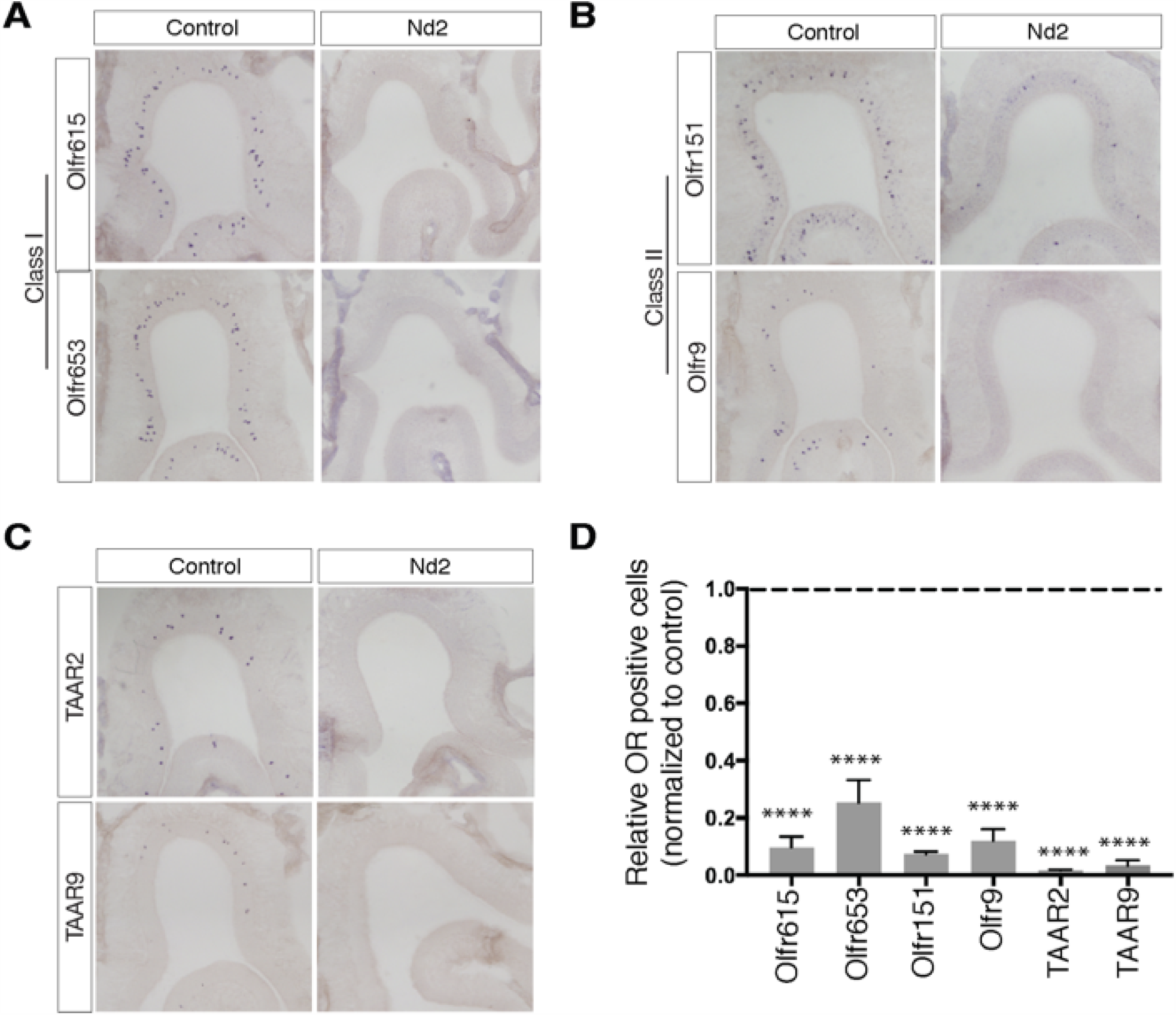
Markedly less OR expression in OSNs in Nd2 mice. **(A)** Representative regions of olfactory epithelium detected by RNA *in situ* hybridization with probes to Class I OR representatives Olfr615 and Olfr653 in Nd2 and a control littermate. **(B)** Representative regions of olfactory epithelium detected by RNA *in situ* hybridization with probes to Class II OR representatives Olfr151 and Olfr9 in Nd2 and a control littermate. **(C)** Representative regions of olfactory epithelium detected by RNA *in situ* hybridization with probes to TAAR receptor representatives TAAR2 and TAAR9 in Nd2 and a control littermate. **(D)** Quantification of total counts of littermate pairs of control and Nd2 mice, revealing a marked reduction of all representative ORs, which is statistically significant (****p ≤ 0.0001; n=3).

**Table S1.** Demographics of patients and healthcare workers testing positive or negative for the COVID-19 virus by RT-PCR. ^a^ Kruskal-Wallis test ^b^ Fisher’s Exact test * negative value indicates that the SARS-CoV-2 RT-PCR test precedes the COVID Smell Test.

| Demographics | COVID<sup>+</sup> patients<br>(n=40) | COVID<sup>-</sup> patients<br>(n=25) | p-value |
| --- | --- | --- | --- |
| Mean age, yrs (SEM) | 35.40 (2.4) | 38.08 (2.7) | 0.205 |
| % Female (N) | 63% (25) | 64% (16) | 1.000 |
| % Reporting smell loss at the time of the smell test | 45% | 8% | 0.002 |
| Average days between RT-PCR test and smell test* | -2.70 (2.3) | -6.56 (3.6) | 0.299 |

